# Comparative Evaluation of Central Venous Oxygen Saturation, Carbon Dioxide Venous Arterial Gradient, and Lactate Levels as Markers of Tissue Perfusion After Cardiac Surgery: A Prospective Exploratory Observational Study

**DOI:** 10.64898/2026.07.04.26357161

**Authors:** Jade Karoline Neves, Vinicius Venturini, Suely Pereira Zeferino, Filomena Regina Barbosa Gomes Galas, Jose Otavio Costa Auler Junior

## Abstract

**Objective:** This study aims to identify which markers of tissue hypoperfusion—specifically lactate levels, central venous oxygen saturation (ScvO_2_), and venous arterial carbon dioxide gradient (CO_2_ gradient)—have the highest sensitivity and specificity in predicting the discharge of postoperative cardiac surgical patients from the ICU within 48 hours. This is an exploratory, hypothesis-generating investigation.

**Methods:** We conducted a prospective observational study involving 100 patients in the Surgical ICU at the Heart Institute of the Hospital das Clínicas, FMUSP, across two collection periods: February to September 2022 and February to June 2023. The study included patients who underwent surgical procedures requiring cardiopulmonary bypass. Comprehensive data on demographic, clinical, and laboratory variables were collected, focusing on tissue hypoperfusion markers measured at ICU admission and again 24 hours post-admission.

**Results:** Among the evaluated markers, ScvO_2_ measured at 24 hours post-admission showed a statistically significant association with ICU discharge (odds ratio = 1.096, 95% confidence interval = 1.020–1.180, p = 0.012). Formal DeLong’s test confirmed ScvO_2_ at 24h had significantly superior discriminatory performance compared to lactate (AUC 0.661 vs. 0.428; Z = 2.889; p = 0.004). In contrast, other markers—ScvO_2_ at admission, lactate, and CO_2_ gap—did not demonstrate significant associations in the exploratory multivariate analysis (p > 0.05).

**Conclusion:** In this exploratory cohort, ScvO_2_ at 24 hours post-admission showed a statistically significant association with early ICU discharge and demonstrated superior discriminatory performance compared to lactate (DeLong p = 0.004). These findings are hypothesis-generating and require prospective validation in larger, adequately powered studies before clinical recommendations can be made.

## INTRODUCTION

Cardiac surgical procedures are frequently accompanied by profound metabolic disturbances arising from alterations in tissue perfusion. These disturbances are particularly relevant during the first 24 hours following admission to the intensive care unit (ICU), a period in which prompt recognition of circulatory inadequacy is crucial to guide appropriate adjustments in oxygen delivery. Such interventions may include the administration of vasoactive agents and/or targeted fluid resuscitation. Beyond conventional cardiovascular assessment using echocardiography, the integration of static and dynamic hemodynamic variables together with biochemical markers plays a central role in directing therapy and improving postoperative outcomes.(1) Markers of tissue perfusion, such as the venous–arterial carbon dioxide difference (CO_2_ gradient), serum lactate concentration, and central venous oxygen saturation, are routinely incorporated into postoperative monitoring protocols. Although the clinical interpretation of the CO_2_ gradient as an indicator of tissue hypoperfusion remains debated, growing evidence suggests that it may detect perfusion abnormalities earlier and with greater sensitivity than traditional markers, including lactate and venous oxygen saturation.(2) An elevated venous–arterial CO_2_ gradient is thought to reflect impaired cellular oxygen utilization, particularly in pathological states characterized by microcirculatory dysfunction or diffusion limitation, such as tissue edema. (3) Measures of venous oxygenation, including central venous oxygen content (CvO_2_) and mixed venous oxygen saturation (SvO_2_), provide insight into the relationship between global oxygen delivery and systemic oxygen consumption. When oxygen delivery fails to meet metabolic requirements, compensatory increases in oxygen extraction occur, leading to a reduction in venous oxygen parameters. Previous investigations have demonstrated significant associations between venous oxygen saturation and key hemodynamic variables, including cardiac output and the venous–arterial CO_2_ gradient, especially in patients with circulatory compromise or in the postoperative setting following major surgical procedures.(4) Serum lactate assessment remains a cornerstone of postoperative monitoring after cardiac surgery and is typically performed in a serial manner. Persistent elevation or delayed clearance of lactate in the early postoperative phase may indicate ongoing tissue hypoperfusion despite apparently adequate systemic hemodynamics and has been associated with prolonged ICU stay. Consequently, lactate trends are widely used to guide therapeutic decisions, with declining levels considered indicative of effective resuscitation and restoration of adequate circulatory function. (5)

This study presents data derived from a prospective observational cohort comprising 100 consecutive patients admitted to a surgical ICU after undergoing cardiac surgery with extracorporeal circulation. The primary objective was to determine which marker—serum lactate, SvO_2_, or the venous–arterial carbon dioxide gradient—demonstrates the highest sensitivity and specificity for predicting discharge from the surgical ICU within 48 hours. As a secondary objective, the study evaluates the ability of the arterial CO_2_ gap to predict postoperative complications during recovery and compares its prognostic performance with established indicators such as lactate and SvO_2_. Ultimately, this investigation seeks to identify the most robust biomarker for early detection of postoperative circulatory dysfunction in patients undergoing cardiac surgery.

## Methods

### Patients and Study Design

This is a prospective observational study involving 100 patients who underwent cardiovascular surgery across two consecutive collection periods: February to September 2022 and February to June 2023. No formal sample size calculation was performed; accordingly, this study is presented as exploratory and hypothesis-generating, and findings should be interpreted with appropriate caution. The sample comprised consecutive eligible patients admitted during two predefined collection periods, constituting a convenience sample appropriate for the exploratory nature of this study. Patient data were collected prospectively from electronic medical records from the time of ICU admission, with information obtained directly from ICU logs and laboratory databases. The following demographic, clinical, and laboratory variables were recorded: age (years), gender, type of surgery (Inferior Left Ventricular Wall Aneurysmectomy, Tricuspid Valve Repair, Myocardial Revascularization, Acute Myocardial Infarction, Ascending Aorta Replacement, Aortic Valve Replacement, Mitral Valve Replacement, Pulmonary Valve Replacement, Mitral Valve Prosthesis Replacement, Tricuspid Valve Replacement, and Mitral Valve Surgery), smoking status, diabetes, stroke, myocardial infarction, hypertension, congestive heart failure, and alcohol use. Patients were excluded if they refused to participate, experienced perioperative stroke, presented with cognitive dysfunction precluding informed consent, or required permanent pacemaker implantation due to conduction system disorders. Four patients had missing arterial PaCO_2_ values at 24 hours and were excluded only from analyses involving PaCO_2_ and CO_2_ gap (n = 96 for these variables); all 100 enrolled patients were included in ScvO_2_ and lactate analyses.

### Interventions and Data Collection

Tissue perfusion markers and hemodynamic variables were assessed at two time points: ICU admission and 24 hours post-admission. The following parameters were recorded at each time point: SvO_2_ (%), lactate (mg/dL), arterial pCO_2_, venous pCO_2_, and CO_2_ gap. Additionally, vasoactive and inotropic drug use was recorded, including dobutamine, adrenaline, and vasopressin. The primary outcome was ICU discharge within 48 hours of admission. Length of stay was calculated based on admission and discharge dates.

### Statistical Analysis

Qualitative variables were summarized as absolute and relative frequencies; quantitative variables were expressed as means, standard deviations, interquartile ranges, and minimum and maximum values. To address the study’s objectives, patients were divided into two groups based on whether or not they were discharged from the ICU within 48 hours, and compared in terms of age, sex, comorbidities, medication use, and perfusion markers measured at admission and 24 hours later. Qualitative variables were analyzed using Pearson’s Chi-squared test or Fisher’s exact test when expected frequencies were less than five; quantitative variables were assessed using Student’s t-test for independent samples. Variables measured at two time points were compared using paired t-tests and illustrated through individual profile plots and mean plots with 95% confidence intervals. Relationships between indices were evaluated using Pearson’s correlation coefficients and scatterplots. This study is subject to potential selection bias inherent to consecutive single-centre sampling, as unmeasured clinical factors may have influenced both marker levels and outcomes. No blinding was applied to data collection or outcome ascertainment. These limitations are further discussed in the relevant section.

### Logistic Regression and Ethical Approval

To assess the influence of tissue hypoperfusion markers on ICU discharge, both simple and multiple logistic regression models were used, with ICU discharge as the dependent variable. The discriminatory power of each marker was evaluated through ROC curves depicting the trade-off between sensitivity and specificity at different thresholds. The significance level was set at 0.05; statistical analyses were performed using JAMOVI software (version 2.2.5) and R (version 4.0.5). The discriminatory performance of the three perfusion markers was formally compared using DeLong’s test for correlated ROC curves, implemented in the pROC package in R. This study was approved by the Institutional Review Board of the Hospital das Clínicas da Faculdade de Medicina da Universidade de São Paulo (CAAE: 52648921.7.0000.0068; Ethics Committee Opinion No. 5.127.618); all patients or their legal representatives provided written informed consent prior to enrollment, and the study was conducted in accordance with the principles of the Declaration of Helsinki (revised Edinburgh, 2000).

## RESULTS

### Demographic Data

All 100 enrolled patients were included in the primary analyses (ScvO_2_ and lactate). Four patients with missing arterial PaCO_2_ data at 24h were excluded only from CO_2_-derived analyses (n = 96 for those variables). All patients completed the 24-hour follow-up, with no losses. The study included 100 patients admitted for cardiovascular surgery (Table 1), with a predominance of males (70%) over females (30%). The mean age was 61.9 ± 11.3 years, with an age range of 21 to 78 years. In terms of medical history, there was a high prevalence of systemic arterial hypertension (78%), congestive heart failure (49%), smoking (46%), diabetes mellitus (26%), previous myocardial infarction (49%), and a history of stroke (6%). Regarding pharmacological support, approximately one-third of patients required dobutamine (36%) and adrenaline (36%) during the perioperative period.

**Table 1.** Baseline patient characteristics and initial clinical data according to ICU discharge within 48 hours.

| Characteristics | Total patients<br>(n = 100) | ICU discharge:<br>Yes (n = 39) | ICU discharge: No<br>(n = 61) | p-value |
| --- | --- | --- | --- | --- |
| **Sex – n (%)** |  |  |  | 0.227 |
| Female | 30 (30.0%) | 9 (23.1%) | 21 (34.4%) |  |
| Male | 70 (70.0%) | 30 (76.9%) | 40 (65.6%) |  |
| **History – n (%)** |  |  |  |  |
| Hypertension (HTN) | 78 (78.0%) | 26 (66.7%) | 52 (85.2%) | **0.029** |
| Heart failure (HF) | 49 (49.0%) | 20 (51.3%) | 29 (47.5%) | 0.715 |
| Previous myocardial infarction (MI) | 49 (49.0%) | 20 (51.3%) | 29 (47.5%) | 0.715 |
| Diabetes | 26 (26.0%) | 9 (23.1%) | 17 (27.9%) | 0.594 |
| Stroke (CVA) | 6 (6.0%) | 2 (5.1%) | 4 (6.6%) | 0.769 |
| Smoking | 46 (46.0%) | 15 (38.5%) | 31 (50.8%) | 0.277 |
| Alcohol consumption | 10 (10.0%) | 3 (7.7%) | 7 (11.5%) | 0.539 |
| **Drug use – n (%)** |  |  |  |  |
| Dobutamine | 95 (95.0%) | 35 (89.7%) | 60 (98.4%) | 0.054 |
| Epinephrine | 36 (36.0%) | 11 (28.2%) | 25 (41.0%) | 0.194 |
| Vasopressin | 2 (2.0%) | 1 (2.6%) | 1 (1.6%) | 0.747 |
| **Markers (Admission)** |  |  |  |  |
| SvO<sub>2</sub> - Mean (SD) | 76.39 (9.21) | 78.16 (9.14) | 78.16 (9.14) | 0.126 |
| Lactate - Median (p25-p75) | 20 (14.0-27.0) | 22 (14.5-29.0) | 20 (14.0-27.0) | 0.518 |
| CO<sub>2</sub> GAP - Mean (SD) | 7.58 (3.46) | 7.56 (3.51) | 7.59 (3.45) | 0.972 |

Notes: n= number of cases; SD = standard deviation; p25-p75 = 25th and 75th percentiles; HTN = hypertension; HF = heart failure; MI = myocardial infarction; CVA = cerebrovascular accident; ScvO<sub>2</sub> = central venous oxygen saturation. Bold values indicate statistical significance (p < 0.05).

Table 2 summarizes the descriptive statistics and temporal comparisons of ScvO_2_, CO_2_ gap, and lactate levels measured at ICU admission and 24 hours thereafter. Statistically significant changes were observed in several parameters over this period. The mean ScvO_2_ value significantly decreased from 76.39 ± 9.21% at admission to 73.92 ± 8.6% after 24 hours (p = 0.021). This population-level decline reflects the expected post-CPB trajectory of elevated systemic oxygen consumption (SIRS, rewarming, increased metabolic demand); within this context, the absolute ScvO_2_ value at 24h — not the direction of change — carries the discriminatory information for ICU discharge (see Discussion). A similar trend was observed for lactate levels, which decreased from 22.86 ± 10.86 mg/dL to 18.59 ± 8.88 mg/dL (p = 0.001). Regarding arterial blood gases, a significant decrease in arterial Pa CO_2_ was observed, from 44.92 ± 6.85 mmHg at admission to 40.63 ± 5.72 mmHg at 24 hours (p < 0.001). Data for arterial PaCO_2_ were unavailable for four patients, and these cases were excluded from the corresponding analysis. Venous Pv CO_2_ also decreased significantly, from 52.50 ± 6.34 mmHg at admission to 48.44 ± 5.01 mmHg after 24 hours (p < 0.001). In contrast, there was no statistically significant change in the CO_2_ gap, which remained relatively stable with mean values of 7.58 mmHg at admission and 7.86 mmHg at 24 hours (p = 0.536). Patients with missing arterial PaCO_2_ data were also excluded from the CO_2_ gap analysis because of incomplete data.

**Table 2.** Patients’ data at admission and after 24 hours.

| Markers | Admission<br>(n= 100) | After 24 hours<br>(n=100) | Mean difference<br>(95% CI) | p-value |
| --- | --- | --- | --- | --- |
| **SvO<sub>2</sub> (%)** |  |  |  |  |
| Mean (SD) | 76.39 (9.21) | 73.92 (8.6) | 2.468 | 0.021 |
| Median (p25-p75) | 76.55 (70.78 - 83.43) | 74.74 (68.07 - 79.90) | (0.374-4.562) |  |
| Minimum-Maximum | 54.10 - 95.10 | 53.10 - 98.00 |  |  |
| **Lactate (mg/dl)** |  |  |  |  |
| Mean (SD) | 22.86 (10.86) | 18.59 (8.88) | 4.270 | 0.001 |
| Median (p25-p75) | 20 (14.00 - 27.00) | 17.00 (8.00 - 68.00) | (1.768 – 6.772) |  |
| Minimum-Maximum | 6.00 - 59.00 | 8.00 - 68.00 |  |  |
| **Arterial pCO<sub>2</sub> (mmHg)** |  | **(n = 96)** |  |  |
| Mean (SD) | 44.92 (6.85) | 40.63 (5.72) | 4.482 | <0.001 |
| Median (p25-p75) | 44.30 (40.55 – 49.75) | 39.60 (37.30 – 43.90) | (2.923 – 6.041) |  |
| Minimum-Maximum | 24.80 – 69.70 | 28.60 – 65.10 |  |  |
| **Venous pCO<sub>2</sub> (mmHg)** |  |  |  |  |
| Mean (SD) | 52.50 (6.34) | 48.44 (5.01) | 4.065 | <0.001 |
| Median (p25-p75) | 52.40 (48.72 – 55.58) | 47.80 (45.48 – 51.10) | (2.610 – 5.520) |  |
| Minimum-Maximum | 33.70 – 74.00 | 38.30 – 68.20 |  |  |
| **CO<sub>2</sub> GAP (mmHg)** |  | **(n = 96)** |  |  |
| Mean (SD) | 7.58 (3.46) | 7.86 (3.71) | -0.3000 | 0.536 |
| Median (p25-p75) | 7.75 (5.10 – 9.80) | 7.15 (5.38 – 9.63) | (-1.260 – 0.660) |  |
| Minimum-Maximum | 0.40 – 18.10 | 1.20 – 26.7 |  |  |

**Notes:** n = number of cases; SD = standard deviation; p25-p75 = 25<sup>th</sup> and 75<sup>th</sup> percentiles

Figure 1 presents grouped boxplots of ScvO_2_, CO_2_ gap, and lactate at ICU admission and 24 hours, stratified by ICU discharge outcome (yes/no), replacing the individual profile plots as suggested by Reviewer B. Notable inter-individual variability and outliers are apparent across all markers, underscoring the complexity of postoperative physiological behavior and the need for cautious interpretation of aggregate results.

**Figure 1.** Grouped boxplots of ScvO_2_, CO_2_ gap, and lactate at ICU admission and 24 hours, stratified by ICU discharge outcome (yes/no). [Figure replaced as per Reviewer B suggestion.]

Table 3 presents the correlation matrix for SvO_2_, lactate, and CO_2_ gap measured at ICU admission and 24 hours post-admission. Most observed correlations were weak, with only two reaching statistical significance. A significant negative correlation was identified between SvO_2_ and CO_2_ gap at admission (R = −0.377, p < 0.001), suggesting that higher SvO_2_ levels were associated with lower CO_2_ gap values. However, this relationship was not sustained after 24 hours (R = −0.078, p = 0.452). Additionally, a significant negative correlation was found between the CO_2_ gap at admission and lactate levels measured after 24 hours (R = −0.238, p = 0.017). In contrast, correlations between identical parameters measured at both time points were not statistically significant for SvO_2_ (R = 0.299, p = 0.002), lactate (R = 0.149, p = 0.139), or CO_2_ gap (R = −0.078, p = 0.451).

**Table 3.**
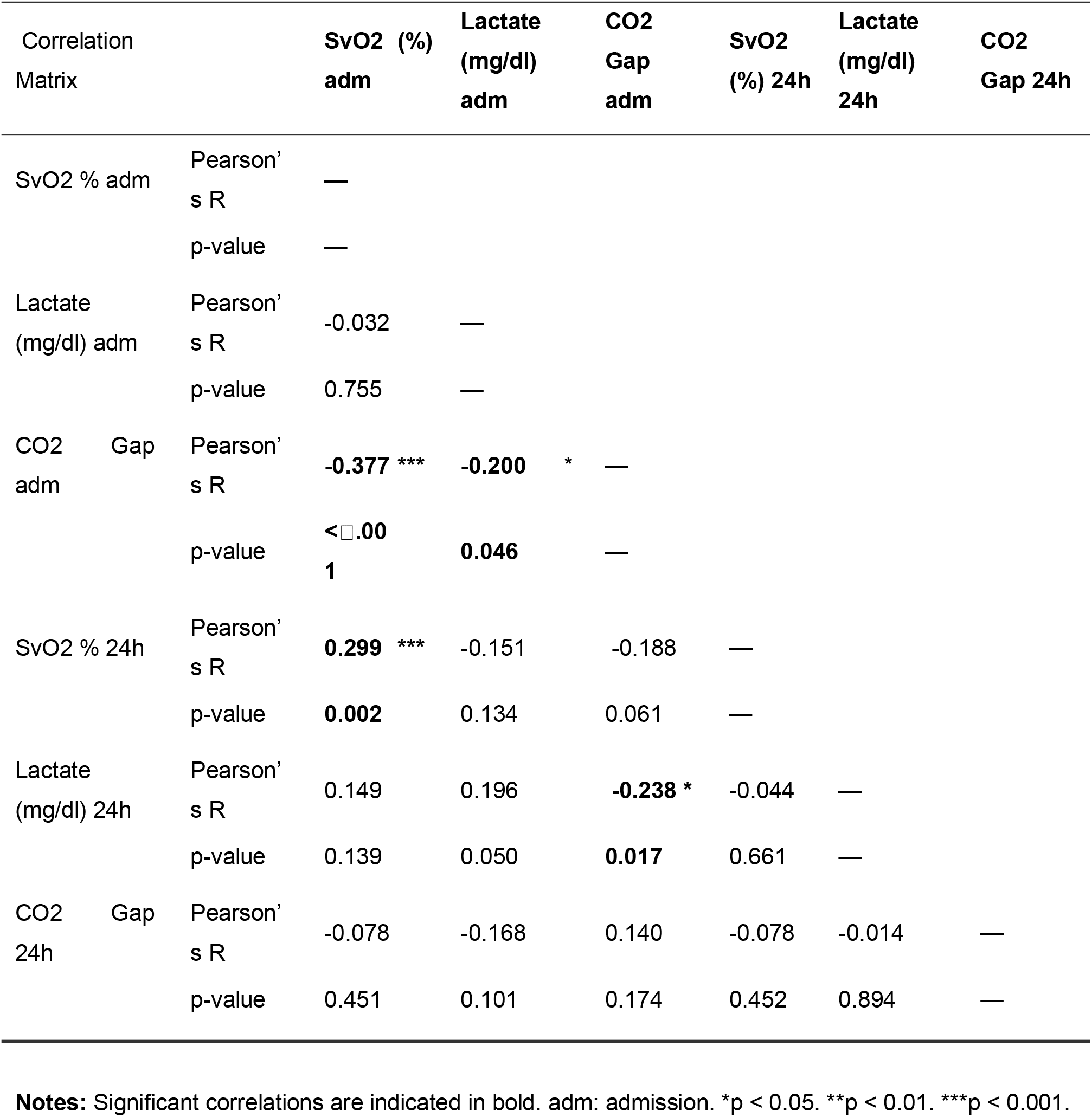
Correlation matrix of SvO2, Lactate, and CO2 GAP variables at admission and 24 hours.

### Comparison between patients with and without ICU discharge within 48 hours

Among the 100 patients included in the study, 39% (n = 39) were discharged from the ICU within 48 hours. The remaining 61 patients (61%) had ICU stays ranging from 2 to 15 days, with most remaining hospitalized for 2 to 3 days. The median ICU length of stay was 3 days (percentile 25 = 2; percentile 75 = 4 days).

Table 4 compares patients discharged from the ICU within 48 hours with those not discharged, based on baseline clinical characteristics and perfusion markers measured at admission and 24 hours post-admission. A statistically significant association was identified between ICU discharge and the presence of systemic arterial hypertension: 26% of patients who were discharged had hypertension, compared to 52% of those who remained in the ICU (p = 0.029). No significant associations were observed for other comorbidities, including congestive heart failure, myocardial infarction, diabetes, stroke, smoking, or alcohol consumption. Regarding pharmacological support, the use of dobutamine demonstrated a trend toward significance, with 35% of patients who received dobutamine being discharged, compared to 60% of those who did not (p = 0.054). Additionally, ScvO_2_ levels measured 24 hours after admission were significantly associated with ICU discharge (p = 0.004).

**Table 4.**
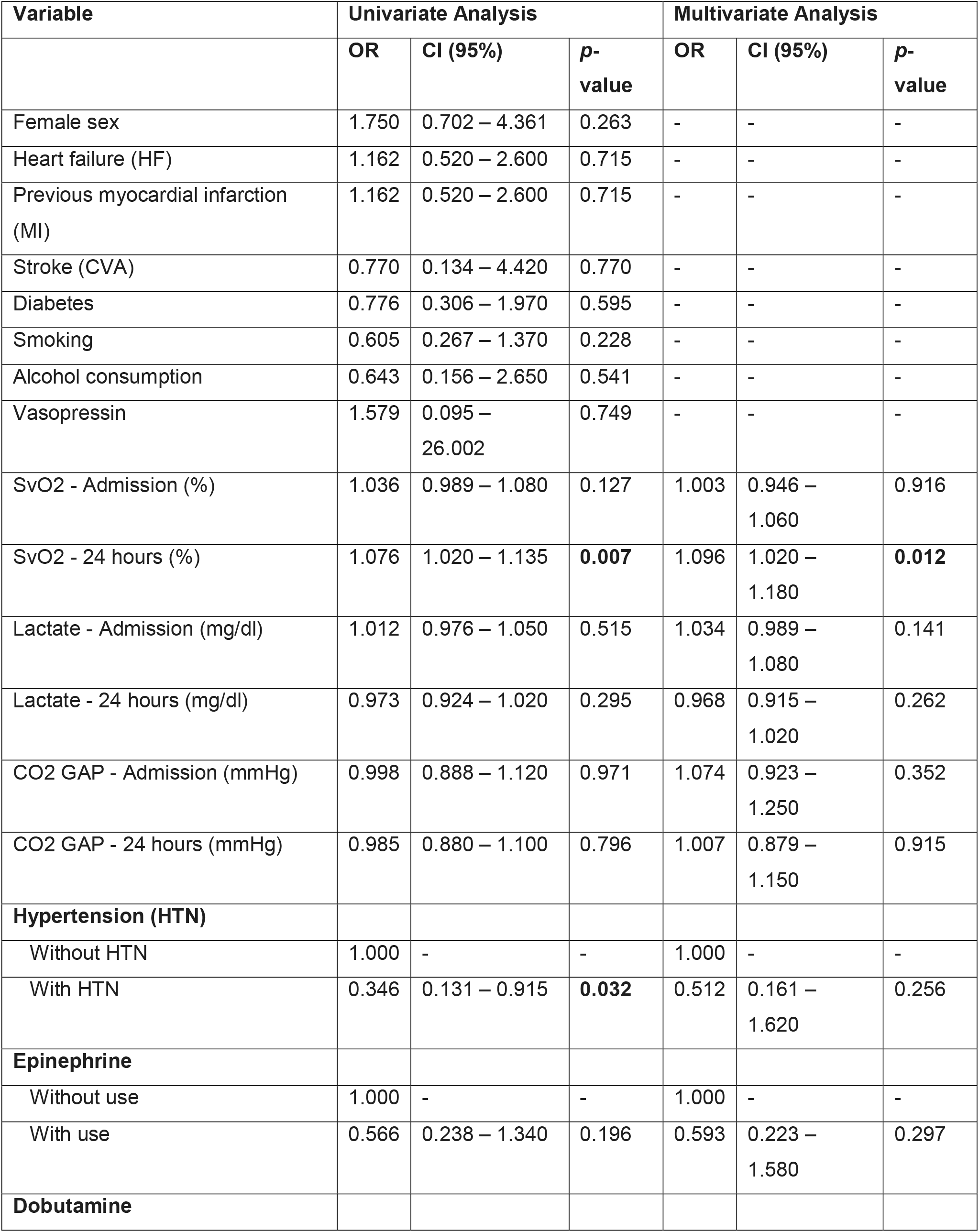
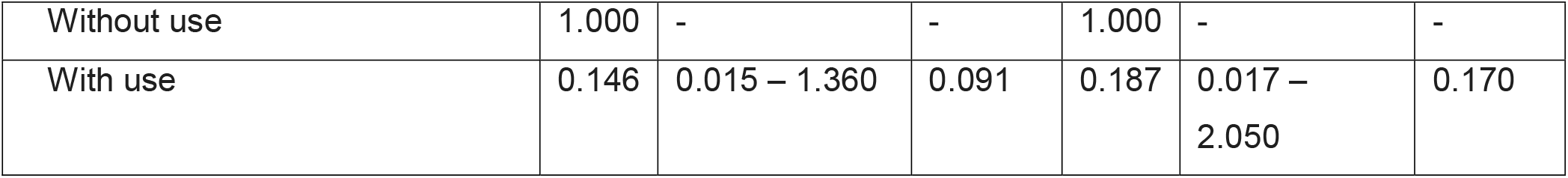
Univariate and multivariate logistic regression analysis of predictors associated with ICU discharge within 48 hours.

| Variable | Univariate Analysis |  |  | Multivariate Analysis |  |  |
| --- | --- | --- | --- | --- | --- | --- |
|  | OR | CI (95%) | p-value | OR | CI (95%) | p-value |
| Female sex | 1.750 | 0.702 – 4.361 | 0.263 | - | - | - |
| Heart failure (HF) | 1.162 | 0.520 – 2.600 | 0.715 | - | - | - |
| Previous myocardial infarction (MI) | 1.162 | 0.520 – 2.600 | 0.715 | - | - | - |
| Stroke (CVA) | 0.770 | 0.134 – 4.420 | 0.770 | - | - | - |
| Diabetes | 0.776 | 0.306 – 1.970 | 0.595 | - | - | - |
| Smoking | 0.605 | 0.267 – 1.370 | 0.228 | - | - | - |
| Alcohol consumption | 0.643 | 0.156 – 2.650 | 0.541 | - | - | - |
| Vasopressin | 1.579 | 0.095 – 26.002 | 0.749 | - | - | - |
| SvO2 - Admission (%) | 1.036 | 0.989 – 1.080 | 0.127 | 1.003 | 0.946 – 1.060 | 0.916 |
| SvO2 - 24 hours (%) | 1.076 | 1.020 – 1.135 | **0.007** | 1.096 | 1.020 – 1.180 | **0.012** |
| Lactate - Admission (mg/dl) | 1.012 | 0.976 – 1.050 | 0.515 | 1.034 | 0.989 – 1.080 | 0.141 |
| Lactate - 24 hours (mg/dl) | 0.973 | 0.924 – 1.020 | 0.295 | 0.968 | 0.915 – 1.020 | 0.262 |
| CO2 GAP - Admission (mmHg) | 0.998 | 0.888 – 1.120 | 0.971 | 1.074 | 0.923 – 1.250 | 0.352 |
| CO2 GAP - 24 hours (mmHg) | 0.985 | 0.880 – 1.100 | 0.796 | 1.007 | 0.879 – 1.150 | 0.915 |
| **Hypertension (HTN)** |  |  |  |  |  |  |
| Without HTN | 1.000 | - | - | 1.000 | - | - |
| With HTN | 0.346 | 0.131 – 0.915 | **0.032** | 0.512 | 0.161 – 1.620 | 0.256 |
| **Epinephrine** |  |  |  |  |  |  |
| Without use | 1.000 | - | - | 1.000 | - | - |
| With use | 0.566 | 0.238 – 1.340 | 0.196 | 0.593 | 0.223 – 1.580 | 0.297 |
| **Dobutamine** |  |  |  |  |  |  |
| Without use | 1.000 | - | - | 1.000 | - | - |
| With use | 0.146 | 0.015 – 1.360 | 0.091 | 0.187 | 0.017 –<br>2.050 | 0.170 |

Results of the simple logistic regression models (univariate analysis) evaluating the impact of clinical and laboratory variables on ICU discharge (Table 5), show that among clinical history variables, the presence of systemic arterial hypertension (SAH), as well as the use of adrenaline and dobutamine, demonstrated statistically significant or borderline associations with ICU discharge outcomes.

The presence of systemic arterial hypertension (SAH) was significantly associated with a lower likelihood of ICU discharge (p = 0.032; OR = 0.345; 95% CI = [0.131 – 0.915]). Similarly, the administration of adrenaline was linked to a reduced probability of discharge, although this association did not reach statistical significance (p = 0.196; OR = 0.33; 95% CI = [0.238 – 1.340]). While the use of dobutamine was not statistically significant, it demonstrated a trend toward a lower chance of ICU discharge (p = 0.091; OR = 0.146; 95% CI = [0.015 – 1.360]).

In an exploratory adjusted multivariate analysis, a logistic regression model was constructed including variables with p-values < 0.2 in the univariate analysis—specifically SAH, adrenaline, and dobutamine—as well as the three key perfusion markers (SvO_2_, lactate, and CO_2_ gap). The results of this model are presented in Table 6.

As shown in Figure 2, only ScvO_2_ measured at 24 hours demonstrated discriminatory capacity for predicting ICU discharge (AUC = 0.661; 95% CI = [0.551–0.772]; p = 0.007). Formal DeLong’s test revealed ScvO_2_ at 24h had significantly superior AUC compared to lactate at 24h (AUC 0.661 vs. 0.428; Z = 2.889; p = 0.004) and a non-significant trend versus CO_2_ gap at 24h (AUC 0.672 vs. 0.520; Z = 1.934; p = 0.053). The remaining indices presented lower AUC values and did not reach statistical significance.

**Figure 2.** ROC curve analysis for ScvO_2_, lactate, and CO_2_ gap with formal DeLong’s test comparison. ScvO_2_ at 24h demonstrated significantly superior AUC vs. lactate at 24h (p = 0.004). CI, confidence interval; AUC, area under the curve; adm, at admission.

## Discussion

In this exploratory single-center study, central venous oxygen saturation (ScvO_2_) measured 24 hours after cardiac surgery showed the strongest association with early ICU discharge among the three markers evaluated, outperforming serum lactate and the venous–arterial carbon dioxide difference. These findings should be interpreted as hypothesis-generating given the modest AUC (0.661), absence of a formal power calculation, and potential unmeasured confounders. ScvO_2_ at 24 hours showed a statistically significant association with early ICU discharge in the exploratory adjusted model. The population-level decline in ScvO_2_ between admission and 24h does not contradict this finding: within the expected post-CPB physiological trajectory — driven by SIRS, rewarming, and elevated metabolic demand — patients who maintained relatively higher absolute ScvO_2_ at 24h demonstrated better preservation of the oxygen delivery-to-consumption ratio, reflecting more efficient hemodynamic recovery. It is the absolute value at 24h, not the direction of change, that carries the discriminatory signal for early ICU discharge.

ScvO_2_, measured via central venous catheter, has long been regarded as a surrogate indicator of systemic oxygen transport, integrating cardiac output, hemoglobin concentration, arterial oxygenation, and tissue oxygen extraction (6). Unlike true mixed venous oxygen saturation (SvO_2_), which requires a pulmonary artery catheter, ScvO_2_ typically overestimates SvO_2_ by approximately 5–8% and reflects predominantly upper-body venous drainage; this physiological distinction should be considered when comparing our findings to prior literature. Large observational studies support its prognostic relevance in cardiac surgery populations. Holm et al. (7) demonstrated that SvO_2_ values below 60% at ICU admission were associated with increased short- and long-term mortality, higher complication rates, and prolonged ICU stay regardless of intraoperative cardiac function, while patients with preserved SvO_2_ exhibited consistently better outcomes. The relationship between venous oxygen saturation and postoperative outcomes has also been explored in the context of occult hypoperfusion: Hu et al. (8) described a high prevalence of patients with reduced ScvO_2_ despite preserved systemic hemodynamics, in whom sustained venous oxygenation impairment combined with hyperlactatemia was associated with longer ICU stays, consistent with the concept that isolated macrohemodynamic stability does not necessarily equate to adequate tissue perfusion. Further complexity is highlighted by Balzer et al. (9), who reported increased mortality at both low and supranormal ScvO_2_ values following cardiac surgery, emphasizing that SvO_2_ interpretation must be contextualized within the broader clinical and physiological framework. In contrast to SvO_2_, lactate levels showed a clear and significant decline during the first 24 postoperative hours, consistent with recovery from perioperative metabolic stress, yet were not associated with ICU discharge. This finding suggests that while lactate is a sensitive marker of global metabolic derangement, it may lack specificity for identifying subtle circulatory inefficiencies that delay readiness for ICU discharge. The prognostic value of lactate following cardiac surgery is well established when elevations are pronounced or persistent (5), and several large studies have demonstrated associations between elevated lactate and mortality, complications, and prolonged ICU stay (11–14). Nevertheless, in the present cohort, lactate kinetics did not discriminate patients suitable for early ICU discharge, underscoring its limitations when used in isolation for this specific outcome. The venous–arterial carbon dioxide gradient similarly failed to demonstrate prognostic relevance: CO_2_ gap values remained elevated but stable over the first 24 hours and were not associated with ICU discharge. Although an inverse correlation between the CO_2_ gap and SvO_2_ was observed at ICU admission, this relationship dissipated over time, suggesting that the CO_2_ gap may reflect early perfusion abnormalities but loses discriminatory capacity as postoperative physiological conditions evolve. Previous investigations have reported conflicting results regarding CO_2_-derived indices (2,6,15–19), and variability between central and mixed venous measurements, as well as postoperative alterations in pulmonary gas exchange, temperature, and regional blood flow distribution, may further confound their interpretation (20).

Receiver operating characteristic analysis supported ScvO_2_ at 24 hours as the most informative variable, with modest discriminative performance (AUC = 0.661; 95% CI = [0.551–0.772]). Formal DeLong’s test confirmed significantly superior discrimination compared to lactate at 24h (AUC 0.661 vs. 0.428; Z = 2.889; p = 0.004); the comparison with CO_2_ gap was borderline (AUC 0.672 vs. 0.520; Z = 1.934; p = 0.053). An AUC of 0.661 indicates modest discrimination and should not support individual clinical decision-making. Lactate and CO_2_ gap exhibited lower accuracy and lacked significant associations with the primary outcome. Notably, ScvO_2_ measured at ICU admission did not predict discharge, highlighting the importance of temporal assessment rather than reliance on isolated early measurements. In addition, a history of systemic arterial hypertension was independently associated with a lower likelihood of ICU discharge, possibly reflecting chronic vascular remodeling, impaired microvascular autoregulation, and reduced physiological reserve, reinforcing the need to integrate biomarker data with baseline clinical characteristics when assessing postoperative recovery trajectories.

The present study has several important limitations. First, the single-center design limits generalizability. Second, no formal sample size calculation was performed; the convenience sample of 100 patients may lack adequate statistical power, and the study must be interpreted as exploratory and hypothesis-generating. Third, venous oxygen saturation was measured via central venous catheter (ScvO_2_), not via pulmonary artery catheter (true mixed SvO_2_); these are physiologically distinct, and findings may not be directly comparable to studies using mixed venous measurements. Fourth, important potential confounders were not prospectively collected, including CPB duration, aortic cross-clamp time, transfusion volumes, STS-PROM, right ventricular function, pulmonary hypertension status, and frailty indices; their absence precludes claims of independent prediction. Fifth, ICU discharge within 48 hours is a soft endpoint subject to institutional influences; harder outcomes (mortality, AKI, low cardiac output, ICU readmission) were not systematically collected. Sixth, the 24-hour assessment time point limits the characterization of ScvO_2_ as a truly early predictive marker. Future studies should incorporate formal sample size calculations, hard clinical endpoints, comprehensive covariate collection, and multicenter designs.

## Conclusion

In this exploratory single-center cohort, ScvO_2_ measured 24 hours after cardiac surgery showed a statistically significant association with early ICU discharge and demonstrated significantly superior discriminatory performance compared to lactate (DeLong p = 0.004), with a borderline trend versus CO_2_ gap (p = 0.053). In contrast, serum lactate and the venous–arterial CO_2_ gradient did not demonstrate significant associations with the primary outcome. However, the modest AUC (0.661), absence of formal power calculation, soft primary endpoint, and unmeasured confounders preclude definitive clinical recommendations. These findings are hypothesis-generating and should be confirmed in prospective, multicenter, adequately powered studies incorporating hard clinical outcomes and comprehensive covariate adjustment.

## Data Availability

De-identified individual participant data are available upon reasonable request to the corresponding author, subject to approval by the Institutional Review Board of the Hospital das Clinicas da FMUSP (CAAE: 52648921.7.0000.0068).

